# Long-term Exposure to Ambient PM_2.5_ and Hospitalizations for Myocardial Infarction among U.S. Residents: A Difference-in-Differences Analysis

**DOI:** 10.1101/2023.03.23.23287669

**Authors:** Yichen Wang, Xinye Qiu, Yaguang Wei, Joel D. Schwartz

## Abstract

**Background:** Air pollution has been recognized as an untraditional risk factor for myocardial infarction (MI). However, the MI risk attributable to long-term exposure to fine particulate matter (PM_2.5_) is unclear, especially in younger populations, and few studies represented the general population.

**Methods:** We applied the difference-in-differences approach to estimate the relationship between annual PM_2.5_ exposure and hospitalizations for MI among U.S. residents and further identified potential susceptible subpopulations. All hospital admissions for MI in ten U.S. states over the period 2002-2016 were obtained from the Healthcare Cost and Utilization Project State Inpatient Database.

**Results:** In total, 1,914,684 MI hospital admissions from 8,106 ZIP codes in ten states from 2002 to 2016 were included in this study. We observed a 1.35% (95% CI: 1.11-1.59%) increase in MI hospitalization rate for 1 μg/m^3^ increase in annual PM_2.5_ exposure. The estimate was robust to adjustment for surface pressure, relative humidity and co-pollutants. In the population with exposure at or below 12 μg/m^3^, there was a larger increment of 2.17% (95% CI: 1.79-2.56%) in hospitalization rate associated with 1 μg/m^3^ increase in PM_2.5_. Young people (0-34 years) and elderly people (≥75 years) were the two most susceptible age groups. Residents living in more densely populated or poorer areas and individuals with comorbidities were observed to be at a greater risk.

**Conclusions:** This study indicates long-term residential exposure to PM_2.5_ could lead to increased risk of MI among U.S. general population. The association persists below current standards.

**Clinical Perspective:** *What is new?:*

- Long-term exposure to PM_2.5_ increased the risk of myocardial infarction in the general U.S. population.
- Young individuals aged 0-34 years had the highest relative risk from long-term exposure to PM_2.5_, and elderly people aged ≥75 years were the second most susceptible to the effects.
- Individuals with iron deficiency anemia, psychosis, and renal failure were more susceptible to the long-term effects of PM_2.5_ on MI.

*What are the clinical implications?:*

- Long-term PM_2.5_ exposure is one of the important modifiable environmental risk factors for myocardial infarction, therefore, air pollution control and behavioral interventions should be taken to prevent the occurrence of myocardial infarction.

## 1. Introduction

Myocardial infarction (MI) is responsible for years of life lost and reduced life expectancy across the globe (Ojha and Dhamoon, 2022). Over the last few decades, the morbidity and mortality burden of MI has declined in developed countries due to improved healthcare systems and preventive actions (Camacho et al., 2022). However, an increasing incidence of MI over time has been reported in younger populations with its underlying reasons unclarified (Arora et al., 2019; Yang et al., 2020).

Over 90% of the risk for acute MI is accounted for by modifiable risk factors (Chadwick Jayaraj et al., 2019). Air pollution is an important driver of MI with a population attributable fraction of 5-7%, (Nawrot et al., 2011). This is similar to well-recognized behavioral factors (e.g., physical exertion, alcohol, and coffee). To date, the epidemiological evidence on the relationship between long-term exposure to ambient fine particulate matter ≤2.5 μm in aerodynamic diameter (PM_2.5_) and MI remains inconclusive. Some studies suggested a positive association (Hartiala et al., 2016; Madrigano et al., 2013), whereas other research did not observe a significant risk associated with PM_2.5_. According to a recent review article, the risk of incident MI could be increased by 8% (95% CI: −1-18%) per 10-μg/m^3^ increase in long-term PM_2.5_ exposure, but the association is evidently weaker as compared to other cardiovascular end points (Alexeeff et al., 2021). In addition, most of the studies used data from cohorts that were not representative of the general population and mostly involving the participants over middle age, such as the EuroTARGET cohort (Gandini et al., 2018) and the Nurses’ Health Study cohort (Puett et al., 2009). Seldom has the effect modification by age been explored in the general population, which, however, is important for understanding the role of air pollution in MI occurrence at earlier ages and potentially different risk profiles among age groups.

Furthermore, concerns are raised in terms of the lack of causal interpretability in the existing literature. In contrast to randomized controlled trials (RCTs) that balance covariates across treatment classes via randomization, observational studies are the most powerful method available to study the health effect of air pollution, since randomization to exposure is not possible. Standard observational studies control for potential confounders in the outcome regression, but are often criticized for the potential for omitted confounders. Several alternative causal modeling approaches have been applied in air pollution epidemiology to attempt to mimic randomized trials (Reich et al., 2021). The generalized propensity score is one example for investigating the health effects of continuous pollution exposure. It can remove the confounding bias by measured covariates and give population marginal effect estimates by balancing the covariates among different exposure levels (Schwartz et al., 2021; Wei et al., 2020). Nevertheless, the propensity score method is not able to control for the unmeasured confounders. The difference-in-differences (DID) is a classical quasi-experimental estimator in econometrics designed to control for unmeasured variables (Ashenfelter and Card, 1985; McEwan, 2010). To date, several studies have applied the DID approach to evaluate the effects of time-varying PM_2.5_ exposure and industrial air pollution on mortality (Leogrande et al., 2019; J. Schwartz et al., 2021; Wang et al., 2016; Yitshak-Sade et al., 2019). This modeling design only examines variations in exposure and outcome within geographic locations (in our case ZIP codes), thereby removing any confounding by measured or unmeasured confounders that differ between ZIP codes. It controls for unmeasured confounders that vary over time similarly in all ZIP codes using indicator variables for each calendar year. Measured covariates are controlled for as usual. Therefore, the model framework provides a robust tool to add control for many unmeasured confounders to traditional observational epidemiology.

Using the DID modeling approach, we investigated the effects of annual ambient PM_2.5_ on the hospital admission rate for MI in residents of all ages across ten states of the United States during 2002-2016. Hospitalization data were accessed from the Health Cost and Utilization Project State Inpatient Databases. Furthermore, we examined whether age could modify the relationships. We also explored the effect modification by community socioeconomic status (SES) indicators and individual comorbidities.

## 2. Methods

### 2.1. Assessment of air pollution

Daily predictions of ambient fine particulate matter (PM_2.5_), nitrogen dioxide (NO_2_), and ozone (O_3_) were derived from an ensemble of machine learning algorithms using geographically weighted regression at a resolution of 1 km × 1 km in the contiguous United States. The prediction model involved critical environmental measures from air monitoring data, meteorological conditions, chemical transport model simulations, land-use features, and satellite remote sensing data of aerosol optical depth, and the model was validated with a 10-fold cross-validation (Di et al., 2020, 2019). We computed the exposure for each ZIP code and each year by averaging the grid cell predictions whose centroids were inside the ZIP code polygons or assigning the nearest grid cell predictions for the ZIP codes that do not have polygon representations, and then linked the exposure data to participants based on their residential ZIP code and admission year. To explore the potential lag effects of air pollution on MI hospitalizations, we used moving averages of air pollution data in the admission year and one year before.

### 2.2. Outcome measurement

The inpatient care records used in this study were obtained from the Healthcare Cost and Utilization Program (HCUP) State Inpatient Database (SID) (HCUP Databases, 2021; Qiu et al., 2023, 2022; Steiner et al., 2002). The database has a 97% completeness of all U.S. community hospital discharges. More details about the inpatient databases and data availability are provided in the Supplement.

We analyzed the hospital admission data in the following listed states and years: Arizona (AZ), Michigan (MI), North Carolina (NC), New York (NY), Rhode Island (RI), Washington (WA), New Jersey (NJ) from 2002 to 2016, Maryland (MD, 2009-2016), Georgia (GA, 2010-2016), and Wisconsin (WI, 2012-2016). The inconsistent inclusion of the study years across these states was based on data availability from the States. The hospitalizations for principal diagnosis of MI were identified with reference to the International Classification of Diseases (ICD) codes (ICD-9: 410; ICD-10: I21).

### 2.3. Covariates

Demographic information for each participant comprising age, sex, race, admission year, state, and ZIP code of residence for that year were obtained from the SIDs. Other covariates included in the models were selected based on our prior knowledge, such as ambient seasonal temperature, and area-level SES and behavioral factors. The SES data at the ZIP code tabulation-area level (ZCTA) were downloaded from the U.S. Census for 2002 Summary File 3 and 2011 Summary File 1 (Bureau, U. S. C., 2011, 2002). They were interpolated annually between census years, and further extrapolated using the American Community Surveys after 2010 (Bureau, U. S. C., 2020). They included the percent of female residents, percent of black residents, median household income, percent of residents having high school education or less, and population density. The percent of residents over the age of 65 years living in poverty was additionally used as a proxy for SES. We defined the ZIP codes where the percent of persons ≥65 years old living in poverty was within the lowest 25^th^ percentile among all areas (i.e., <7%) as low-level poverty areas and other ZIP codes as poorer areas. We also linked in the county-level yearly percentage of residents who ever smoked and mean body mass index (BMI) from the Centers for Disease Control and Prevention (CDC) Behavioral Risk Factor Surveillance System (BRFSS) (Bureau, U. S. C., 2020), which were converted to ZCTA-level based on the county and updated annually. Ambient daily surface temperature and daily average surface pressure at a 12 km × 12 km resolution were obtained from the National Aeronautics and Space Administration’s Land Data Assimilation System (NLDAS-2). In light of the seasonal effect of temperature on cardiovascular health (Shi et al., 2015; Stewart et al., 2017), we generated two metrics of temperature, i.e., the average temperature in the warm months (April-September) and the average temperature in the cold months (October-March). The ambient annual average level of daily maximum relative humidity at a 4 km × 4 km resolution was obtained from the gridMET dataset (Abatzoglou, 2013). Temperature, surface pressure, and relative humidity data were later aggregated to ZIP codes and years.

### 2.4. Comorbidity data

In this study, we considered several coexisting medical conditions that are likely to impact the vulnerability of MI patients exposed to PM_2.5_ in the secondary analysis, which included: hypertension (Yang et al., 2022), diabetes mellitus (DM), chronic obstructive pulmonary disease (COPD) (Croft et al., 2022; Rich et al., 2010; Wynands et al., 2022), renal failure (Bo et al., 2021), iron deficiency anemias (IDA) (Pereira and Sarnak, 2003), obesity (Weichenthal et al., 2014), peripheral vascular disorders (PVD) (Bauersachs et al., 2019; Gwon et al., 2021), depression (Gładka et al., 2018), other neurological disorders (Shi et al., 2020), and psychoses (Larsen and Christenfeld, 2009; Qiu et al., 2023, 2022). The comorbidity data was assigned to the MI inpatients using the AHRQ comorbidity software and stored in the Disease Severity Measures file as a part of SIDs. The comorbidity data are available from 2005 to 2015. The identification of comorbidities is based on International Classification of Diseases, 9th revision, Clinical Modification (ICD-9-CM) diagnoses and the Diagnosis-Related Group (DRG) in effect on the discharge date.

### 2.5. Statistical analysis

We used the DID approach to estimate the relationship between long-term exposure to ambient PM_2.5_ and the incidence of hospital admissions for MI. The analysis was limited to the ZIP code-year combinations with a population of more than 100 to reduce the noise from low-population areas and increase the analytical power (Qiu et al., 2022). First, we calculated the annual aggregated counts of hospital admissions for MI for each ZIP code, year, and age group (0-34, 35-54, 55-64, 65-74, ≥75 years). We fitted a quasi-Poisson regression to account for the overdispersion of MI hospitalization counts. The equation is given below:

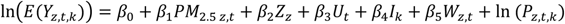

where *Y_z,t,k_* represents the aggregated count of hospitalizations for MI in ZIP code *z*, year *t* and age group *k*; *PM*_2.5 *z,t*_ represents the mean ambient PM_2.5_ concentration for the same stratum of ZIP code *z* and year *t*; *Z_z_* is a dummy variable for ZIP code which captures all time-invariant or slowly changing variables that vary across ZIP code areas, measured or unmeasured; *U_t_* is a dummy variable for year *t* which represents the time-varying variables whose temporal variation is similar across ZIP code areas; *I_k_* is a dummy variable for age groups; *W_z,t_* represents the variables that may vary differently over time across ZIP code areas; ln (*P_z,t,k_*) is an offset term that represents the natural log of the population in ZIP code *z*, year *t* and age group *k*.

The essence of the DID method is to account for all differences across ZIP codes using indicator variables, and only examine the within ZIP code variation in exposure and outcome. This controls for all slowly changing covariates, measured or unmeasured, that vary across ZIP codes. Omitted confounders that vary similarly over time across ZIP codes are controlled using indicator variables for year. Time-varying confounders that vary over time differently across ZIP codes must be measured and controlled as usual. More statistical details can be found elsewhere (Wang et al., 2016).

In our DID model, the spatial confounding was controlled for by fitting individual intercepts for each ZIP code. The time trends were controlled by using an indicator for each year. We controlled for time-varying estimates of average warm-season and cold-season temperatures, percentage of female residents, percentage of black residents, median household income, percent of education level under high school, population density, percentage of residents who ever smoked, percentage aged ≥65 years old living in poverty, and mean BMI in this paper. We assumed no other covariates that are both correlated with exposure and outcome can change differentially over time across ZIP codes were left out other than these adjusted variables. In the ecological design, the individual-level risk factors that displayed temporal variabilities were unrelated to the exposure and therefore would not confound the association. Therefore, the estimates given by the model can reflect the changes in MI morbidity attributable to the changes in ambient PM_2.5_.

To test the robustness of the effect estimate, we further controlled for surface pressure and relative humidity, respectively. We also constructed two- and multi-pollutant models to control for potential confounding by NO_2_, and O_3_. Moreover, we explored the variation in the effect estimates across age subgroups by including an interaction term for an indicator of age subgroups and the PM_2.5_ term. We additionally examined the potential effect modification by some typical area-level SES indicators that could illustrate the degree of urbanity, i.e., poverty level, medium household income, population density, and education level.

We performed two secondary analyses. First, we restricted the analysis to the residents who lived in the areas with annual exposure consistently at or below the annual standard of 12 µg/m^3^ set by National Ambient Air Quality Standards (NAAQS) over the study period. For the above analysis, the percent change in hospitalization rate for MI and their 95% confidence intervals (CIs) per μg/m^3^ increase in long-term average PM_2.5_ level were reported. Second, we employed the case-only analysis to examine whether the following individual-level comorbidities could be effect modifiers: hypertension, DM, COPD, renal failure, IDA, obesity, PVD, depression, other neurological disorders, and psychoses. The case-only method was suggested by Armstrong (2003) to study modification by effectively time-invariant individual factors in research of time-varying environmental factors to identify the susceptible groups. For each comorbidity type, we performed a logistic regression model of the ambient PM_2.5_ on the comorbidity status adjusted for the time trend among the MI inpatients. The exposure-modifier interaction odds ratio (OR) and 95% CIs were reported.

All analyses were performed using R software version 4.1.2. A two-sided p-value <0.05 was considered statistically significant.

## 3. Results

### 3.1. Characteristics of community conditions and MI hospitalizations

In total, we included 1,914,684 hospital admissions for MI from 8,106 ZIP codes (with a population over 100) in 10 U.S. states from 2002 to 2016. The air pollution, temperature, demographic and SES characteristics averaged over the study period in these ZIP codes are presented in **Table 1**. Over the study period, the average long-term annual PM_2.5_ concentration was 8.9±2.6 μg/m^3^ among all the ZIP code areas. The average temperatures in the warm season and in the cold season were 19.0±3.7 and 4.7±4.8 °C, respectively. On average, female residents, ever smokers, or residents who received high school education or less accounted for about half of the ZIP code-level population. In addition, there was evidence of spatial heterogeneity in several community-level demographic and SES covariates. For example, the population density differed substantially across ZIP codes with its 10^th^ and 90^th^ percentiles of 9 and 2,062 persons/mi^2^, respectively. The 10^th^ and 90^th^ percentiles in the percentage of the low-education population (≤high school) ranged between 25.2% and 63.6%.

**Table 1.** Distribution of ZIP code-level annual PM_2.5_ and covariates from 2002 through 2016.

| ZIP code-level covariates* | Mean (SD) | Percentile |  |  |  |  |
| --- | --- | --- | --- | --- | --- | --- |
|  |  | 10th | 25th | 50th | 75th | 90th |
| Annual PM<sub>2.5</sub> (µg/m<sup>3</sup>) | 8.9 (2.6) | 5.4 | 7.2 | 8.9 | 10.5 | 12.4 |
| Annual NO<sub>2</sub> (ppb) | 17.6 (10.0) | 7.4 | 10.1 | 14.8 | 23.3 | 31.9 |
| Annual O<sub>3</sub> (ppb) | 39.0 (3.9) | 34.7 | 37.3 | 38.9 | 40.5 | 44.0 |
| Warm season temperature (°C) | 19.0 (3.7) | 15.0 | 16.3 | 18.1 | 21.6 | 23.9 |
| Cold season temperature (°C) | 4.7 (4.8) | -0.8 | 0.9 | 4.3 | 7.8 | 11.1 |
| Surface pressure (Pa) | 95,668 (2,980) | 94,187 | 94,267 | 94,330 | 98,118 | 100,097 |
| Relative humidity (%) | 84.8 (10.6) | 74.9 | 83.8 | 87.6 | 90.4 | 92.9 |
| Percent female (%) | 50.3 (3.6) | 47.7 | 49.4 | 50.6 | 51.8 | 53.0 |
| Percent black (%) | 9.9 (17.1) | 0.2 | 0.5 | 2.2 | 10.5 | 31.6 |
| Median household income (\$) | 54,044<br>(23,198) | 32,364 | 39,049 | 48,130 | 63,069 | 83,717 |
| Percent $\geq 65$ below poverty | 13.2 (8.5) | 4.33 | 7.06 | 11.53 | 17.06 | 24.17 |
| Population density<br>(persons/mi<sup>2</sup>) | 1,109 (3,866) | 9 | 23 | 96 | 753 | 2,062 |
| Percent education ≤ high<br>school | 46.0 (14.6) | 25.2 | 35.9 | 47.7 | 56.9 | 63.6 |
| Ever smokers (%) | 48.0 (7.5) | 39.0 | 43.3 | 47.6 | 52.5 | 57.4 |
| Mean BMI (kg/m<sup>2</sup>) | 27.6 (1.1) | 26.3 | 26.9 | 27.5 | 28.2 | 29.0 |

\* Air pollutant concentration data were the moving averages of the year and the previous year.

**Table 2** shows the total and age subgroup-specific number of hospitalizations for MI and the corresponding average hospital admission rate among the included ZIP codes during the study period. Among all 1,914,684 MI cases, the highest hospital admission rate was occurring in patients aged at least 75 years old, which accounted for 35.98% of total MI hospitalizations. The proportions of MI hospital admissions in 45-54, 55-64, and 65-74 age groups were lower than that in the oldest group, being 14.20, 21.92, and 22.51%, respectively. In contrast, the age group aged 0-34 years old only included 0.79% of total hospitalizations for MI. The annual admission rate in the general population across all ZIP code areas shows an increasing pattern with age group: The highest ZIP code-level annual hospitalization rate for MI of 10.62‰ was observed in the oldest age group (≥75 years), while lower hospitalization rates were seen in younger age groups.

**Table 2.**
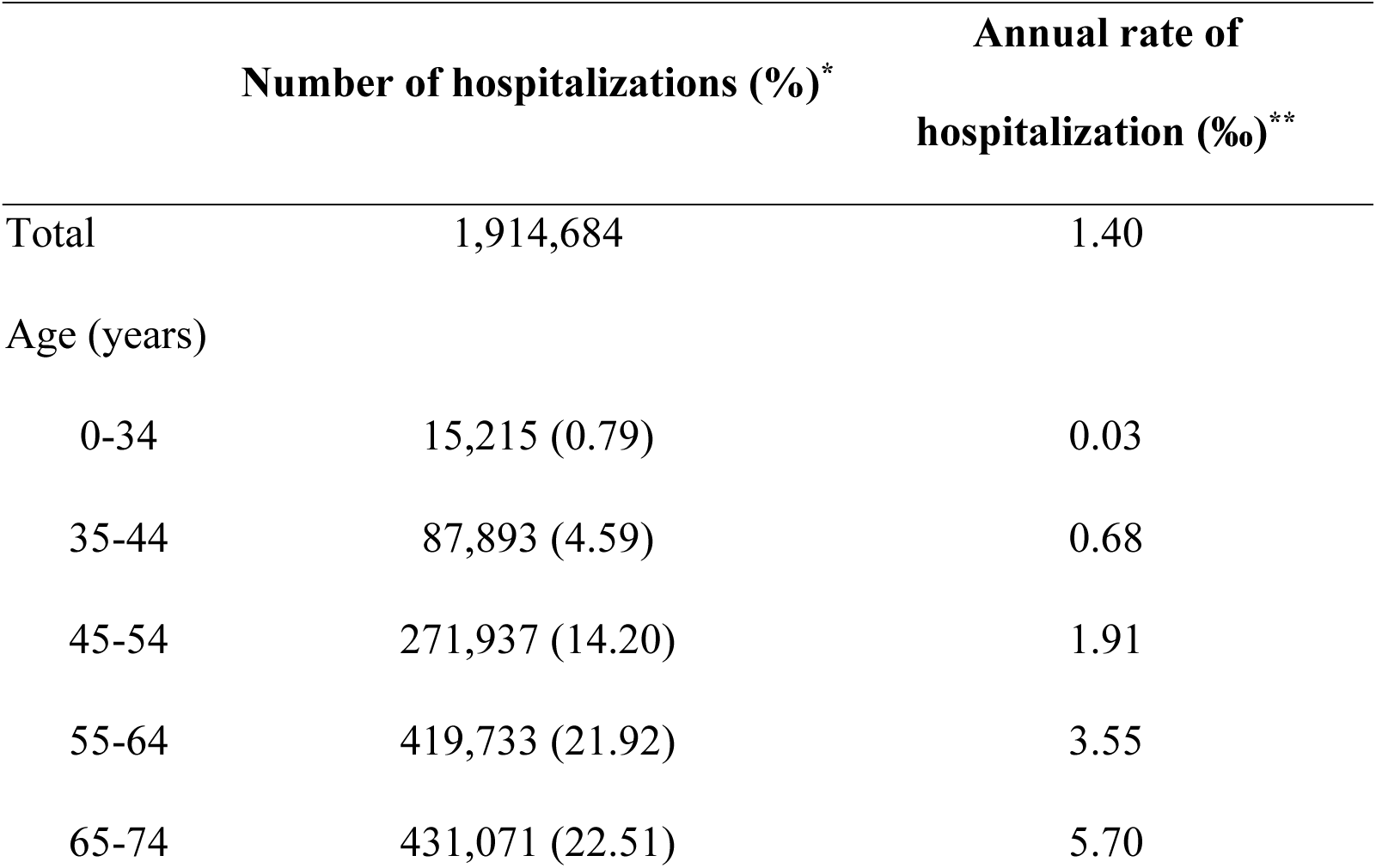
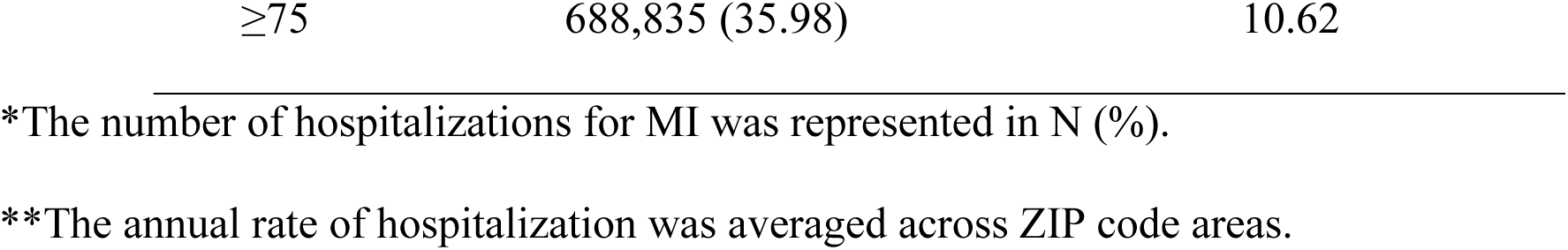
Summary of total and age-specific hospitalizations for MI from 2002 through 2016.

### 3.2. Main effects of long-term PM_2.5_ exposures on MI risk

**Table 3** shows the estimated effects of PM_2.5_ on MI risk in the general population using different models. If the model assumptions are met, these are causal estimates. Overall, we observed a 1.35% (95% CI: 1.11-1.59%) increase in the hospitalization rate for every 1-μg/m^3^ increase in PM_2.5_ exposure. The estimate was stable after further adjusting for surface pressure or relative humidity. In the model further adjusted for NO_2_ or O_3_, the adverse effects of PM_2.5_ on the risk of MI were slightly attenuated, which corresponded to 1.23% (95% CI: 0.99-1.47%) and 1.18% (95% CI: 0.94-1.42%) increase in hospital admission rate associated with 1 μg/m^3^ increase in PM_2.5_, respectively. In the model adjusted for NO_2_ and O_3_ simultaneously, the effect estimate was similar and remained significant. Moreover, we also restricted the main analysis to the ZIP codes where the annual PM_2.5_ exposures were always below 12 μg/m^3^ over the study period. A higher increment of 2.17% (95% CI: 1.79-2.56%) in hospitalization rate for MI was observed per 1-μg/m^3^ increase in the low-level PM_2.5_ exposures.

**Table 3.**
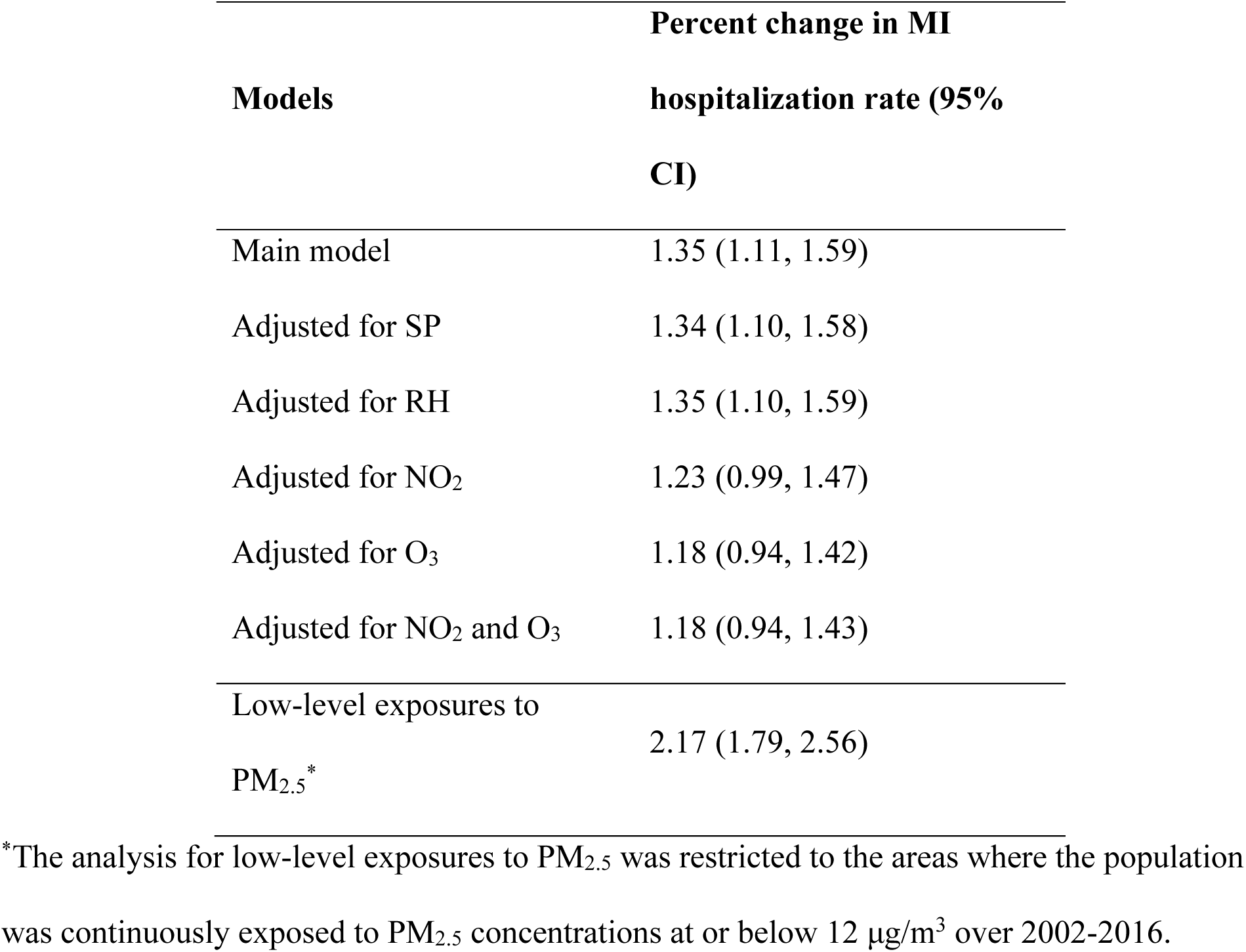
Percent change in hospitalization rate for MI per 1-μg/m^3^ increase in long-term exposure to PM_2.5_ in the main model, in models adjusted for surface pressure (SP) or relative humidity (RH), in models adjusted for NO_2_ and/or O_3_, and in the low-level exposure analysis.

### 3.3. Heterogeneity of susceptibilities among age subgroups

**Figure 1** and **Table S1** show the variation in estimates for annual PM_2.5_ and hospitalization for MI across six age subgroups. In detail, the subgroups aged 0-34 and ≥75 years appeared to have the highest relative risks for chronic exposures to PM_2.5_. For every 1-μg/m^3^ increase in annual PM_2.5_, there was a 2.40% (95% CI: 1.62-3.20%) and a 1.94% (95% CI: 1.69-2.20%) increase in the hospitalization rate in 0-34 years and ≥75 age subgroups, respectively. Every 1-μg/m^3^ increase in PM_2.5_ was associated with an increment of 1.32% (95% CI: 0.93-1.70%) in the hospitalization rate for MI among the subpopulation aged 35-44 years old. In contrast, the lowest relative risks for PM_2.5_ were observed in the subpopulations with ages 45-54, 55-64, and 65-74 years.

**Figure 1.**
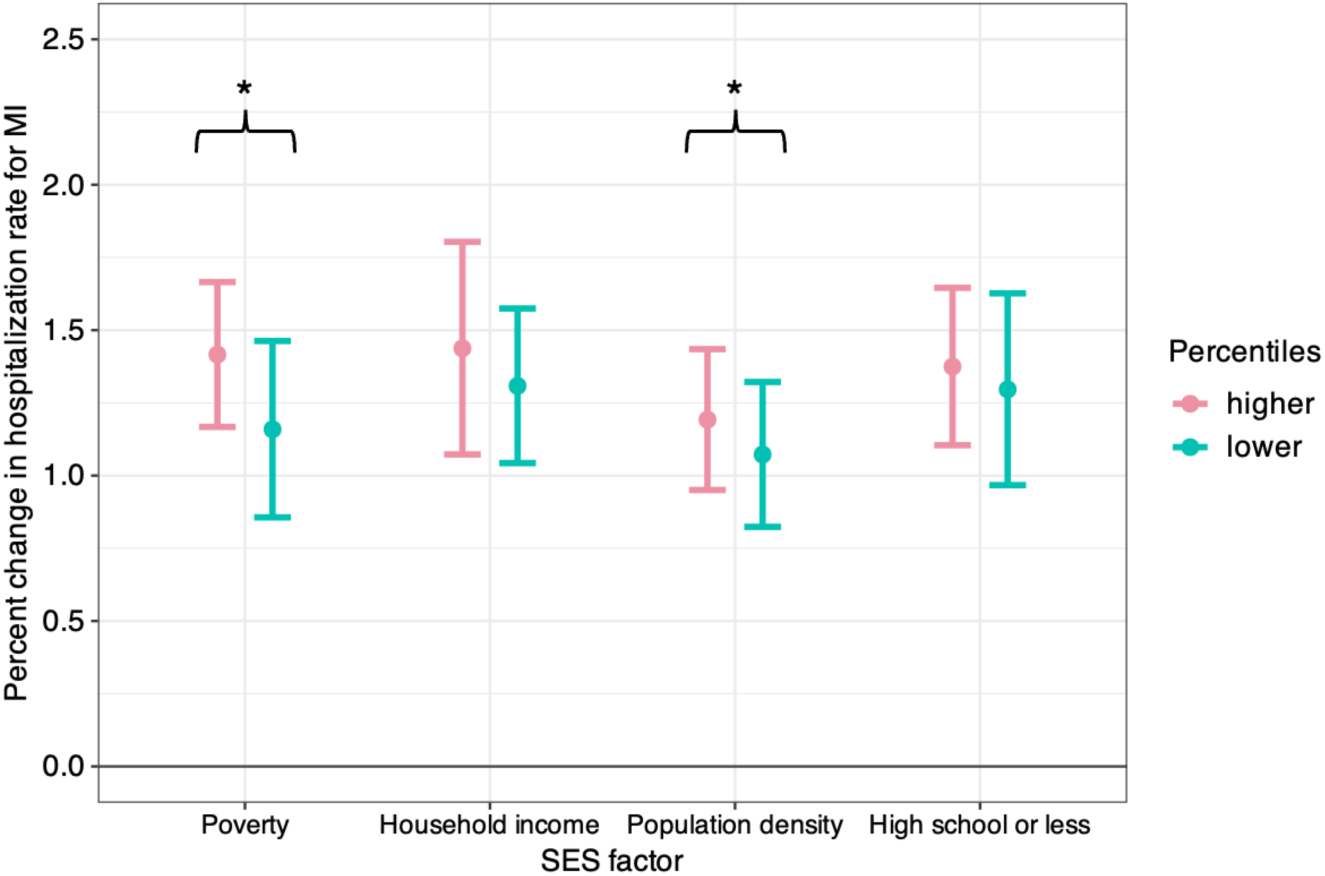
Percent change in hospitalization rate for MI per 1-μg/m^3^ increase in long-term exposure to PM_2.5_ across different age subgroups.

### 3.4 Effect modifications by area-level SES covariates

**Figure 2** and **Table S2** illustrate the potential effect modification by several area-level SES covariates on the relationship between PM_2.5_ and the risk of MI. We found significant effect modification by area-level poverty (*p* _interaction_=0.0497). Specifically, the population living in the poorer areas was more susceptible to long-term PM_2.5_ exposures as compared to those living in lower-poverty areas. In addition, population density significantly modified the association (*p* _interaction_<0.0001). The result suggests that people living in more populated geographical areas tended to have a higher risk of MI attributed to long-term PM_2.5_ exposures. However, the association was not modified by either median household income (*p* _interaction_=0.5190) or education level (*p* _interaction_=0.6640), despite a tendency of the higher risk occurring in the population with lower median household income and lower education level.

**Figure 2.**
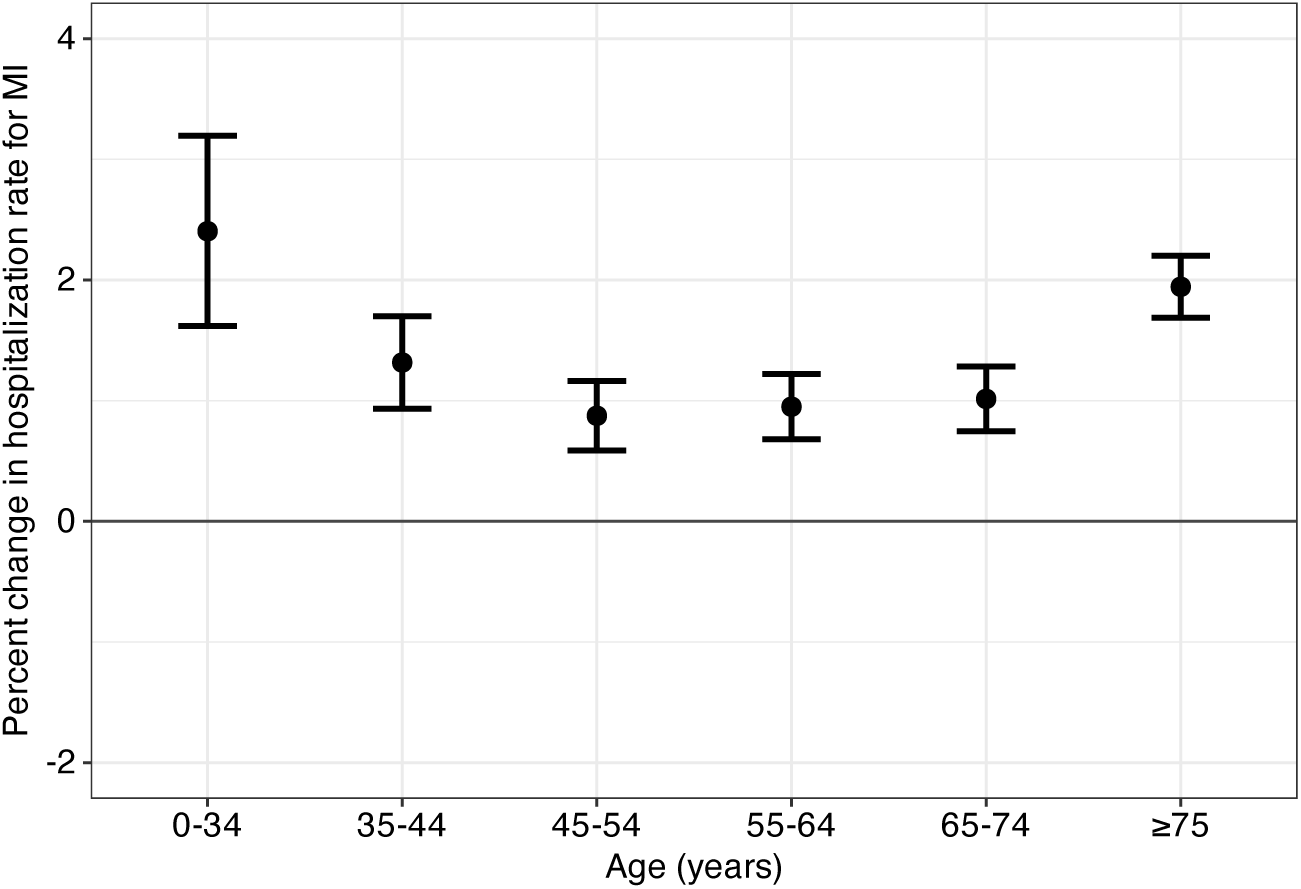
Percent change in hospitalization rate for MI per 1-μg/m^3^ increase in long-term exposure to PM_2.5_ at the different percentiles of potential SES modifiers. * indicates the statistical significance of the modifier (*p* _interaction_<0.05). For the percent of residents ≥65 years old living in poverty, the data below the 25^th^ percentile (i.e., <7% percent; defined as low-level poverty areas) compared to the data at or above the 25^th^ percentile (i.e., ≥7% percent; defined as poorer areas). For median household income, population density, and percent education level ≤high school, the effects of PM_2.5_ were compared between the 10^th^ and 90^th^ of the modifier level.

### 3.5. Effect modifications by individual comorbidities

We also investigated whether the association between long-term PM_2.5_ and hospitalizations for MI could differ by individual-level comorbidities using the data from 2005 to 2015. The results of the case-only analysis are shown in **Figure 3** and **Table S3**. The risk of MI due to chronic exposures to PM_2.5_ was significantly higher in individuals with most of the common comorbidities than in those without it: hypertension, DM, COPD, renal failure, IDA, obesity, PVD, other neurological disorders, and psychoses. The strongest modifying effect was observed in individuals with comorbid IDA (*p* _interaction_<0.0001). The odds of hospitalizations for MI was increased by 5.6% (95% CI: 5.3-5.8%) for each 1-μg/m^3^ increase in PM_2.5_ associated with comorbid psychoses. However, we found that having depression may decrease the susceptibility to PM_2.5_ exposures (OR=0.995; 95% CI: 0.991-0.998).

**Figure 3.**
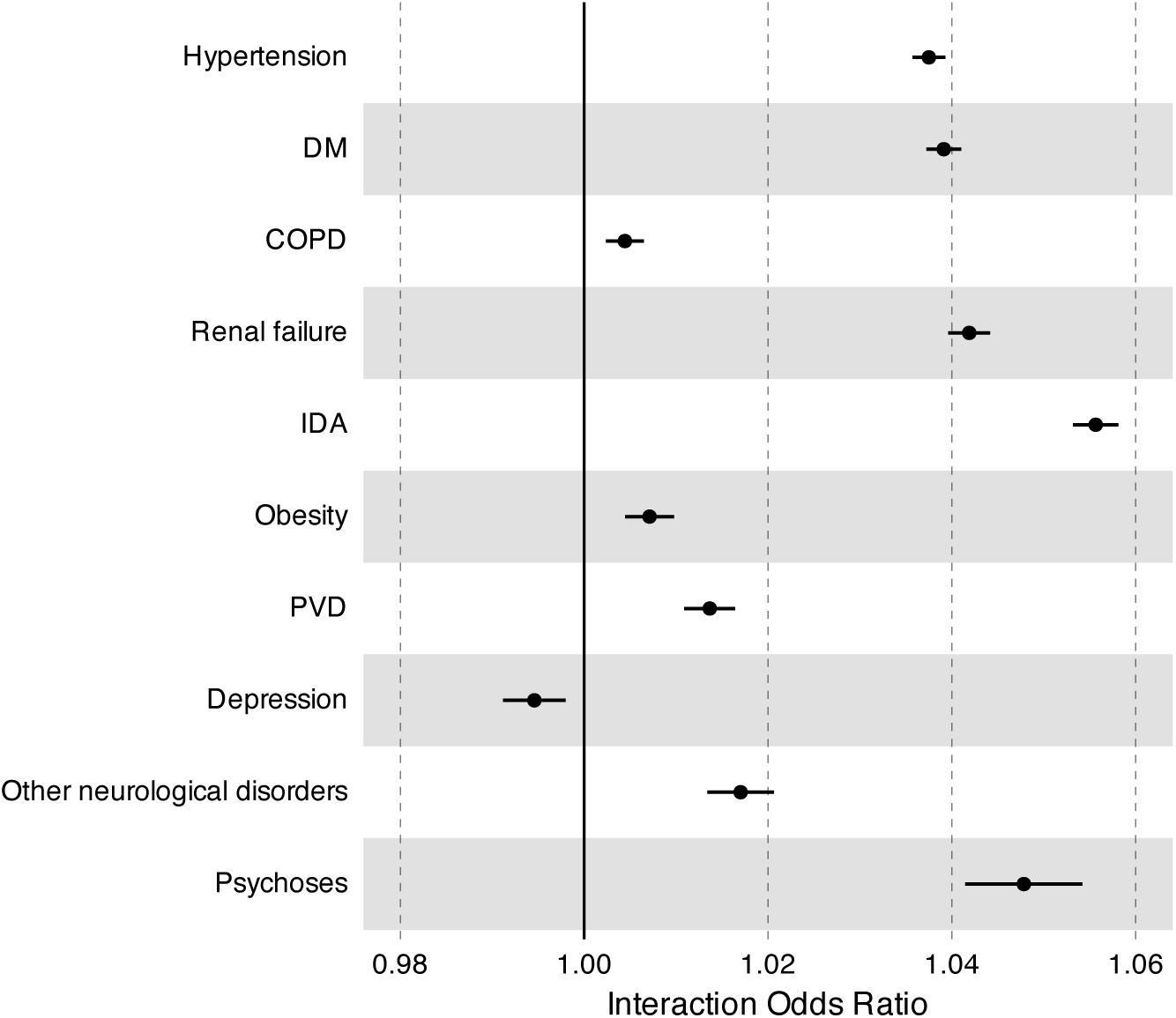
Interaction Odds Ratio (OR) for hospitalization for MI per μg/m^3^ increase in long-term exposure to PM_2.5_ comparing individuals with and without comorbidities. DM, diabetes mellitus; COPD, chronic obstructive pulmonary disease; IDA, iron deficiency anemia; PVD, peripheral vascular disorders.

## 4. Discussion

In this study, we used hospital inpatient records from HCUP SIDs covering all admissions in ten states in the U.S. and performed a DID analysis to estimate the relationship between long-term exposure to ambient PM_2.5_ and the risk of hospitalizations for MI. In the entire population, we found that the incidence of MI would increase by 1.35% (95% CI: 1.11-1.59%) for every 1-μg/m^3^ increase in long-term residential PM_2.5_ exposure. The effect was robust to adjustment for surface pressure, relative humidity and co-pollutants (i.e., NO_2_ and/or O_3_). Effects remained and were stronger when limited to locations that never exceeded U.S. EPA’s air quality standards, were larger in poorer neighborhoods, and were increased in persons with chronic conditions most of which are more common in minorities. A key aspect of this analysis is that the population studied is the total population of those states, and not the selected sample of most cohort studies. This makes the findings more generally applicable and the coefficients more useful for calculating attributable risks. The findings that people with iron deficiency anemia, psychosis, and renal failure were more susceptible to the effects of PM_2.5_ on MI are novel, and the other effect modification (hypertension, diabetes, COPD) findings add to a thin literature.

Less attention has been paid to the long-term effects of PM_2.5_ as compared to their acute effects (Cai et al., 2016; Zhu et al., 2021); however, an additional 10% risk is estimated for long-term exposure than for short-term exposures to PM_2.5_, partially due to deteriorated cardiometabolic conditions over time (Rajagopalan et al., 2018). Therefore, our research supplements the limited evidence in this regard. The risk of hospitalization for MI was slightly attenuated when NO_2_ and/or O_3_ were accounted for as confounding factors. It is important to note that NO_2_ and O_3_ are associated with certain components of PM_2.5_. In particular, NO_2_ is associated with secondary organic particles and O_3_ with secondary inorganic (sulfate and nitrate) particles. Hence controlling for them may remove the effects of important PM components. Indeed a personal exposure study reported that ambient NO_2_ and O_3_ were not associated with personal exposure to NO_2_ and O_3_ but were associated with personal exposure to PM_2.5_ (Valli, 2001). This suggests that three pollutant models risk biasing downward the PM_2.5_ effect as the gases may not be confounders. This change may also be due to the potential positive association for NO_2_ with MI (Roswall et al., 2017). The possible confounding by O_3_ might be explained by its potential to amplify cardiovascular effects (Malik et al., 2019; Weichenthal et al., 2017), which, however, remains unclear because of its complex dynamics and interaction with other typical air pollutants that may vary with seasons (Ito et al., 2007; Mustafić et al., 2012).

Moreover, we observed that the increased risk was stronger at or below annual PM_2.5_ concentrations of 12 μg/m^3^ which is an indication of a curvilinear concentration-response function (CRF). Such a CRF has been noted previously. For example, Vodonos et al. (2018) in a meta-analysis of 54 cohort studies reported a similar shape in the association of both all-cause mortality and cardiovascular mortality. The Global Exposure Mortality Model study reported a similar finding, with a steeper slope for ischemic heart disease mortality below 12 µg/m^3^ (Burnett et al., 2018). Possible reasons for such a pattern include adaptation, different chemical compositions of particles in locations with lower exposure, a different ratio of surface area to mass in higher concentration locations where particles tend to be larger due to agglomeration, etc. Overall, our results highlight that there is a significant health threat even under low-level long-term exposures and that pollution control measures should be prioritized to protect public health.

Our main finding is in accordance with some recent evidence. For example, Danesh Yazdi et al. (2019) observed a 2.6% (95% CI: 2.4-2.8%) increase in the hazard of first hospital admission for MI associated with each 1 μg/m³ increase in average annual PM_2.5_ concentration among Medicare participants over 65 years or older in seven southeastern states in the U.S. A study targeted the entire adult population in Ontario, Canada reported an ∼3% increase in the risk of MI incidence per μg/m^3^ increase in long-term exposure to PM_2.5_ (Chen et al., 2020). Among 118,229 individuals in a large-scale and population-based Chinese cohort, an increment of 10 μg/m³ increase in PM_2.5_ was associated with a higher risk of acute MI (HR=1.28; 95% CI, 1.19-1.39) (Li et al., 2020). Hystad et al. (2020) found that a hazard ratio of 1.03 (95% CI: 1.00-1.05) per 10 μg/m³ increase in long-term ambient PM_2.5_ and a population-attributable fraction of 8.4% (95% CI: 0.0-15.4%) for MI in the Prospective Urban and Rural Epidemiology cohort comprising 157,436 adults from 21 countries. Nevertheless, some researchers reported null associations of long-term PM_2.5_ with MI (Cesaroni et al., 2014; Lipsett et al., 2011; Miller et al., 2007). The nationwide Danish Nurse Cohort Study reported a higher risk of fatal MI induced by PM_2.5_, but did not suggest any association with overall incident MI (Cramer et al., 2020). Aside from the statistical approaches and study period, the differential magnitudes of the effect among the studies may be due to differences in demographic structure, prevailing concentrations, since the association appears curvilinear but linear models were fit, physiological characteristics (e.g., medical history, comorbidities), or SES factors (e.g., poverty, healthcare accessibility) of the study population. Diverse sources and constituents of ambient PM_2.5_ across different regions can also explain the difference in the toxicity and the triggering of MI (Rich et al., 2013).

Multiple pathways by which PM_2.5_ can affect the myocardium have been heavily elaborated, which are linked to oxidative stress, endothelial dysfunction, systemic immunity response, systemic inflammatory response, and autonomic nervous system injuries (Langrish et al., 2012; Rajagopalan et al., 2018; Wang et al., 2013; Wang et al., 2016; Zhao et al., 2013); these physiological responses could further contribute to the development of atherosclerosis, the widely believed main trigger and underlying disease process of MI onset (Palasubramaniam et al., 2019). MI progression could be accelerated by disturbed cardiac mitochondrial function and dynamics followed by impaired redox metabolism and inflammation in the lung due to chronic exposure to PM_2.5_ inhalation (Marchini et al., 2022). More pathological evidence of specific molecular signaling pathways is also emerging. For example, toxicological experiments revealed that the down-regulation of microRNA-205 induced by PM_2.5_ could stimulate myocardial inflammation and cardiac dysfunction (Feng et al., 2020). Hu et al. (2021) demonstrated that NLRP3 inflammasome activation is responsible for PM_2.5_-induced endothelial cell dysfunction. These uncovered biological mechanisms well support our findings.

We provide novel evidence into the health disparities across age groups in terms of the relationship under investigation. Our evidence of high sensitivity in the oldest age group is in line with prior studies (Bai et al., 2019; Li et al., 2020). It could result from reduced physiological, metabolic and compensatory processes, as well as the heavier health burden of cardiopulmonary disorders in the elderly (Shumake et al., 2013). It might be also attributed to declined antioxidant defense ability of the respiratory tract lining fluid in the elderly population (Kelly et al., 2003). We also found a surprisingly high susceptibility in young individuals, which has been seldom explored or discussed before. A similar finding was suggested in a Canadian study where the researchers observed a strong association of MI incidence in relation to PM_2.5_ between 35-44 years-old adults and 75-85 years-old adults (Bai et al., 2019). In comparison, we extended the scope to a large sample size of multiple age groups and found that the susceptibility in individuals aged 0-34 years was higher than that in the middle-aged groups and slightly higher than that in those aged ≥75. It is important to note that the baseline risk in the younger individuals was two orders of magnitude lower than in the oldest category, so a high relative risk is still a small attributable risk. These findings might be partially due to the longer time spent outdoors by younger people than by elder people. It’s also possible that a potentially immature immune system in adolescents may increase their vulnerability (Frcpc et al., 2006). More importantly, while the traditional risk factors for MI such as underlying atherosclerosis are less common in younger populations, their unique risk profile is linked to behavioral and psychosocial factors (e.g., recreational drug use) which may add to the risk of cardiovascular events (Gulati et al., 2020). Abundant evidence concludes cocaine use is a strong risk factor for MI (DeFilippis et al., 2018; Phillips et al., 2009; Schwartz et al., 2010). These risk factors may convey susceptibility to these individuals to the inflammatory response induced by PM_2.5_. On the other hand, if cocaine usage is correlated with PM_2.5_ exposure, it may be a confounder.

In addition, we observed that population living in poorer areas or densely populated areas may be more susceptible to the risk of MI attributed to chronic exposures to PM_2.5_. The evidence of differential susceptibility to exposure in low-income areas, combined with the already established higher exposure levels and poorer healthcare accessibility in socioeconomically disadvantaged areas (Colmer et al., 2020; Wang et al., 2017) identifies this as a key environmental justice issue. Although increased susceptibility to PM_2.5_-related health impacts accompanied by SES disadvantage has been well recognized (Bravo et al., 2016; Chi et al., 2016; Xu et al., 2020), the effect modification by neighboring SES factors in the association of MI risk with long-term PM_2.5_ exposures is little discussed, and present conclusions on this topic are mixed. Several studies did not identify significant differences across levels of neighborhood-level sociodemographic characteristics (Hystad et al., 2020; Madrigano et al., 2013; Weaver et al., 2019). In contrast, the evidence of effect modification by annual household income was suggested in long-term adult residents of Ontario (Bai et al., 2019), which differed from our study. Our present finding can provide a novel understanding of potential susceptible subpopulations and facilitate clarification of social drivers for substantial geographic disparities in MI across the U.S. (Camacho et al., 2022; Yu et al., 2021).

In line with prior epidemiological studies, we identified a statistically elevated risk of MI associated with long-term PM_2.5_ exposure among patients with diabetes than those without (Cramer et al., 2020; Hart et al., 2015; McGuinn et al., 2016). We also found that hypertension, COPD, renal failure, IDA, obesity, PVD, and unfavorable neurological conditions may increase the vulnerability to the risk of MI in relation to PM_2.5_, which has not been widely appreciated or documented in the past. Higher insulin resistance and oxidative stress may enhance the alteration of ventricular repolarization and systemic inflammation triggered by PM_2.5_ and thus exacerbate myocardial vulnerability to arrhythmias (Schneider et al., 2010), which might provide a possible explanation for our conclusion. The strongest effect of IDA may be due to stimulated oxidative stress, decreased blood supply, myocardial hypoxia, and the close interplay of the outcomes (Inserte et al., 2021; Padda et al., 2021). Additionally, we assume the intake of psychotropic medications may exacerbate the susceptibility since they are known to cause undesirable cardiometabolic effects (Abosi et al., 2018; Mohamed et al., 2019). A much lower chance of adherence to cardiovascular disease medication and healthcare-seeking behaviors to prevent physical illness among many psychiatric patients (especially those with severe mental illness) might also contribute to an increased risk of MI events (Druss et al., 2001; Newcomer and Hennekens, 2007). However, comorbid depression, which showed a protective effect, remains unclear. Further investigations on the specific impacts and mechanisms of comorbidities are warranted.

There are multiple strengths in the present study. To begin with, we used high coverage (97%) medical records across multiple U.S. states over a long period, which enabled the statistical power to be sufficient and expanded the generalizability of our findings. Furthermore, in the DID modeling framework, we eliminated confounding by variables that change slowly over time and minimized the confounding by the variables that showed parallel variations in time across area units, thus overcoming the limitations of data availability by design. We also fully adjusted for multiple area-level demographic, SES, and behavioral covariates to relax the assumptions. Additionally, in light of underappreciated cardiovascular risk among younger individuals, we performed the analysis with fine divisions of age and revealed the strong susceptibility in young people. Additionally, we determined multiple common individual comorbid conditions as potential susceptibility factors. With this investigation, we aimed to raise the public health concern towards sensitive groups and implement more targeted preventative measures both in pollution control and healthcare services.

However, some limitations should also be acknowledged. First, despite high resolution of the air pollution predictions, the PM_2.5_ estimates only represent the ambient exposure level within a specific ZIP code area and year but were not based on personal exposures which are influenced by activity patterns, air exchange rates, second-hand tobacco smoke, and other factors that lead to them differing, often substantially, from neighborhood exposure. However, Weisskopf and Webster (2017) have pointed out that personal exposure is correlated with many potential confounders (e.g. stress while driving) that are not associated with outdoor concentration, and hence outdoor exposures can act as instrumental variables for the personal exposure. Further, the National Human Activity Pattern Survey in the U.S. reported that U.S. adults spent 69% of their time at home and 8% of the time immediately outside their home (Klepeis et al., 2001), making it reasonable to use neighborhood ambient pollution to capture their exposure to ambient pollution. Nevertheless, our model clearly does not capture personal exposure. While there is likely a considerable difference between the personal exposure of individuals living in a neighborhood and the ambient concentration in that neighborhood, the difference between the personal and ambient is likely to be mostly Berksonian exposure error since it represents individual variation around a group mean. As such, it should not produce much bias in the effect estimates in an epidemiology study. A recent simulation study showed that even in the presence of substantial Berksonian exposure error, little bias would be found in an epidemiology study (Wei et al., 2022). Second, the DID approach should be better understood as a more robust approach to confounding control that addresses some omitted confounders rather than as an approach that addresses all of them. However, since we used indicator variables for every ZIP code, our analysis is only examining year-to-year exposure within ZIP code, and hence ZIP code differences between e.g. high and low SES ZIP codes are controlled by design. Within ZIP code, we have controlled for time-varying SES variables, but we acknowledge we were not able to control for all. For example, we did not have data on occupation. However, since the DID approach eliminates any contrasts between ZIP codes, and the exposure assigned to all individuals within a ZIP code is the same, it is unlikely that such factors could be associated with exposure. Third, differential exposure histories and other risk factors such as diets across areas are overlooked because of the unknown human migration patterns in this study, which may have resulted in a wrong recognition of the true cause of MI hospitalizations. Finally, we aggregated the hospital admissions by strata defined by ZIP code, year, and age, therefore, potentially low numbers in some age subpopulation in less populated geographical areas could add noise to the analysis.

## 5. Conclusions

In this study, we found that long-term exposures to ambient PM_2.5_ increased the risk of MI hospitalization. The relationship remained significant after controlling for surface pressure, relative humidity and concurrent exposures to NO_2_ and O_3_. A more pronounced adverse association was observed at or below the annual PM_2.5_ limit enacted by NAQQS. In addition, we observed higher susceptibility to the PM_2.5_-related MI risk among young people and elderly people, residents living in poorer or highly populated areas, and individuals with a comorbid burden. Our study highlights the roles of air pollution being one important environmental risk factor for MI, gives insights into the underlying susceptible subpopulations, and informs the future strategies to control air pollution and mitigate related health disparities.

## Data Availability

The authors do not have permission to share data.

## Acknowledgments

We want to offer special thanks to U.S. Agency for Healthcare Research and Quality Health Cost and Utilization Project (HCUP) for giving us access to the State Inpatient Databases (SIDs) to conduct this study.

## Supplementary Materials

### Data S1. Supplemental Methods

HCUP data are developed and managed through a Federal-State-Industry partnership sponsored by the Agency for Healthcare Research and Quality. By providing uniformly formatted datasets for multiple states, the data are aimed to facilitate health services research and improve health care delivery. The SIDs in HCUP data contain the inpatient discharge records from community hospitals in 49 participating states, which account for about 97% of all U.S. community hospital discharges. The coverage of SIDs reaches all patients regardless of payer, including individuals covered by Medicare, Medicaid, or private insurance, and those uninsured.

In the present study, we included the SIDs for the following ten states that were allowed by our current funding budget for research use from HCUP: Arizona (AZ), Michigan (MI), North Carolina (NC), New York (NY), Rhode Island (RI), Washington (WA), New Jersey (NJ), Maryland (MD), Georgia (GA), and Wisconsin (WI). For GA, we only have access to the data after 2010. For WI, we only have access to the data after 2012. For MD, residential address ZIP code information is only available after 2008 and admission year information is only available after 2009. For the other included states, we have the complete records from 2002 to 2016.

**Table S1.** Percent change in hospitalization rate for MI per 1-μg/m^3^ increase in long-term exposure to PM_2.5_ across different age groups.

| Age (years) | % Change (95% CI) |
| --- | --- |
| *p* for trend* | <0.001 |
| 0-34 | 2.40 (1.62, 3.20) |
| 35-44 | 1.32 (0.93, 1.70) |
| 45-54 | 0.87 (0.59, 1.16) |
| 55-64 | 0.95 (0.68, 1.22) |
| 65-74 | 1.01 (0.75, 1.28) |
| $\geq 75$ | 1.94 (1.69, 2.20) |

\**p* for age trend was estimated from the adjusted model using age as a continuous term.

**Table S2.** Percent change in hospitalization rate for MI per 1-μg/m^3^ increase in long-term exposure to PM_2.5_ across different levels of SES factors.

| SES factors | 10th percentile | 90th percentile | *p* for interaction |
| --- | --- | --- | --- |
| Poverty | 1.16 (0.86, 1.46) | 1.42 (1.17, 1.67) | 0.0497 |
| Household income | 1.31 (1.04, 1.57) | 1.44 (1.07, 1.80) | 0.5190 |
| Population density | 1.07 (0.82, 1.32) | 1.19 (0.95, 1.43) | <0.0001 |
| High school or less | 1.30 (0.97, 1.63) | 1.37 (1.10, 1.65) | 0.6640 |

**Table S3.**
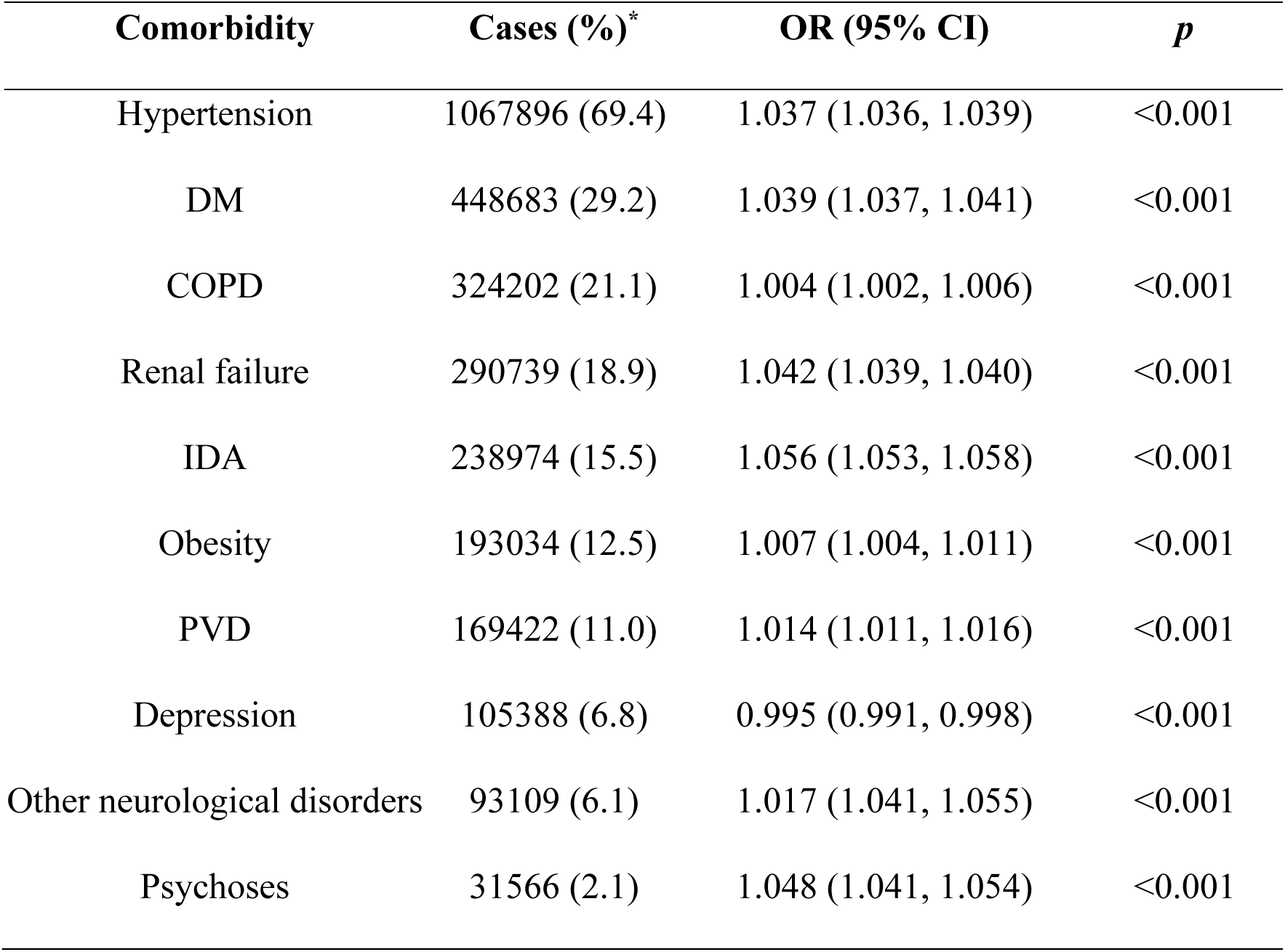
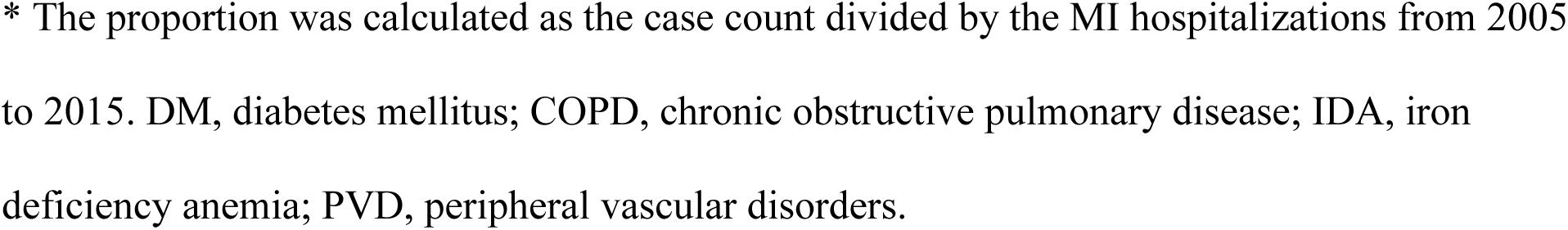
Interaction Odds Ratio (OR) for hospitalization for MI per 1-μg/m^3^ increase in long-term exposure to PM_2.5_ comparing individuals with and without comorbidities.

